# Investigating the Impact of Segregation and Integration on Infection Dynamics: Analysis of a Cholera Model in a Two-Population System

**DOI:** 10.1101/2024.05.01.24306689

**Authors:** Omar El Deeb, Antoine Matar

## Abstract

We present a novel dynamic model designed to depict Cholera outbreaks within a two-population framework featuring two environmental reservoirs. The model is designed to emulate the impact of segregation or integration between two populations on the transmission of the disease and infections throughout the entire community, both with and without non-medicinal interventions. This is achieved by allowing infectious individuals to interact with the reservoir of the alternate population at different levels of suppression, in addition to their regular interaction with their own reservoir.

We find out that increased suppression of cross community interaction reduces the number of infections in the overall population as well as in the population with less contamination and contact. Additionally, we predict significant delays in the occurrence of peak infections, affording public health authorities crucial time for intervention. Lowering cross-immunity interactions also leads to a decrease in bacterial concentrations in environmental reservoirs. Finally, we demonstrate that non-medicinal interventions, including sanitation and water purification, would significantly reduce and delay infections, providing a valuable time frame for implementing additional medicinal measures.

## 1 Introduction

Cholera is a bacterial infection caused by the Vibrio cholerae bacterium, and it is primarily transmitted through contaminated water and food. Cholera epidemics can be dangerous and potentially life-threatening, especially in areas with inadequate sanitation and limited access to clean water [1]. The severity of cholera epidemics depends on various factors, including the local infrastructure, healthcare system, and the ability to implement effective public health measures. Scientists have approximated that annually, there are between 1.3 and 4.0 million instances of cholera, resulting in 21,000 to 143,000 fatalities globally attributable to the disease [2].

It has a long and impactful history, with documented cases dating back centuries [3, 4]. The first cholera pandemic emerged in the early 19th century, originating in the Indian subcontinent and spreading globally through trade routes and military movements. The causative agent, Vibrio cholerae, was identified by the pioneering work of John Snow in the mid-1800s, who linked the disease to contaminated water sources. Subsequent pandemics occurred in waves, affecting diverse regions and populations. Advances in understanding the bacterium, improvements in sanitation, and the development of oral rehydration therapy have contributed to better managing and controlling cholera. Despite these efforts, outbreaks continue to pose challenges, particularly in areas with inadequate access to clean water and sanitation. Cholera remains a significant public health concern, underscoring the ongoing importance of global efforts to prevent, detect, and respond to this infectious disease [5, 6, 7, 8].

The disease manifests with distinct symptoms, primarily characterized by severe diarrhea and vomiting. This intense loss of fluids can lead to dehydration, a critical aspect of the disease. Individuals may also experience nausea and abdominal cramps. In severe cases, cholera can progress rapidly, causing extreme thirst, lethargy, and a rapid heart rate. Without prompt and adequate re-hydration, the symptoms can escalate, potentially leading to shock and, in the absence of proper medical intervention, even death. The severity of symptoms can vary, and some infected individuals may exhibit milder or asymptomatic cases, contributing to the challenge of detecting and containing the disease in affected populations.

Cholera has the potential to rapidly spread in regions characterized by inadequate sanitation, crowded living conditions, and poor hygiene practices, primarily due to contamination of water sources. The symptoms, including severe diarrhea and vomiting, can swiftly lead to dehydration and, without timely re-hydration, pose a fatal threat, especially to vulnerable populations such as young children and the elderly. Beyond the individual health implications, cholera outbreaks significantly impact communities, causing social and economic disruptions by straining healthcare systems, overwhelming medical facilities, and contributing to elevated mortality rates. The disease’s prevention and effective management hinge on enhancing surveillance systems, ensuring access to clean water, proper sanitation facilities, and timely medical care, with oral re-hydration therapy playing a crucial role in saving lives [9]. While cholera is commonly associated with developing countries and poor sanitation, it can also emerge in areas affected by natural disasters or conflicts, and the risk of global spread persists, especially with increased international travel.

Mathematical modeling plays a crucial role in understanding and predicting the spread of infectious diseases. By employing mathematical frameworks, researchers can simulate and analyze the complex dynamics of disease transmission within populations. These models take into account various factors such as population size, demographics, contact patterns, and the effectiveness of interventions. They provide valuable insights into the potential trajectories of an outbreak, helping public health officials make informed decisions about implementing control measures. Mathematical models also allow for the exploration of different scenarios, enabling researchers to assess the impact of interventions and identify strategies to mitigate the spread of the disease [10, 11]. Moreover, these models contribute to our understanding of the underlying mechanisms driving epidemics, aiding in the development of more effective public health strategies and interventions [12, 13, 14, 15]. The interdisciplinary nature of mathematical modeling in infectious disease research highlights its significance in addressing global health challenges [16].

The transmission dynamics of infectious diseases have undergone comprehensive studies through the application of the Susceptible-Infectious-Recovered (SIR) compartmental model, initially introduced by Kermack and McKendrick [17], and its subsequent refinements and adaptations. Over the years, this model has been employed to analyze a spectrum of diseases, ranging from historical pandemics such as the Spanish flu to endemic illnesses like Cholera, Malaria, and Pneumonia, as well as contemporary challenges like the seasonal flu and the ongoing COVID-19 pandemic [18, 19, 20, 21, 22, 23, 24, 25, 26]. The versatility of these compartmental models has been demonstrated in their ability to effectively simulate and predict the trajectories of disease spread, accounting for variables such as mitigation measures, sanitation practices, social distancing initiatives, and vaccination campaigns. By leveraging these models, researchers and public health officials gain valuable insights into the potential impact of interventions, enabling more informed decision-making to mitigate the consequences of infectious diseases [27, 28, 29, 30, 31].

The use of SIR compartmental models has proven to be invaluable in the field of cholera modeling, providing a robust framework for understanding and predicting the dynamics of cholera outbreaks. Cholera often leads to rapid and widespread transmission, making it crucial to develop effective strategies for its control and prevention [32]. In the context of cholera, individuals in the susceptible compartment are those who are at risk of contracting the disease, while infectious individuals have been exposed to the bacterium and can transmit the infection. The recovered compartment includes individuals who have either survived the infection and gained immunity or succumbed to the disease. This model captures the essential dynamics of cholera transmission, allowing researchers and public health officials to estimate key parameters such as the basic reproduction number *R*_0_, which represents the average number of secondary infections generated by a single infectious individual in a completely susceptible population. The SIR model also helps in assessing the impact of interventions such as vaccination, water sanitation measures, and improved hygiene practices on controlling cholera outbreaks. By incorporating real-world data into the model, researchers can tailor interventions to specific settings and populations, ultimately contributing to more effective and targeted cholera prevention strategies. Additionally, the SIR model’s adaptability allows for the exploration of various scenarios and the identification of optimal control measures to mitigate the impact of cholera on communities and prevent its further spread.

Extensive research exists on modeling cholera, incorporating an environmental reservoir that typically signifies a community’s water source, housing the Vibrio Cholerae bacteria responsible for infections in individuals within the community. Interactions with the reservoir result in individuals shedding more bacteria into it through various mechanisms [33, 34, 35, 36, 37]. While some models account for person-to-person transmission of the infection [38], this aspect remains a secondary factor in disease dynamics compared to water and environmental contamination. The majority of models in the literature concentrate on a singular population interacting with a sole reservoir, with certain studies examining clusters of populations interacting with a shared reservoir [39, 40, 41].

In this study, we highlight a specific scenario that holds significant importance in certain contexts. Our primary focus is on a society consisting of two populations, each associated with distinct environmental reservoirs that exhibit varying degrees of interaction. An illustrative example involves a guest community coexisting with a host community, each possessing its own water resources and experiencing different levels of segregation or integration. In a completely segregated society, the two populations and their reservoirs operate independently, whereas in a fully integrated society, they function as a unified entity. If a cholera infection spreads within one population, health authorities must formulate policies to mitigate the disease and alleviate the burden on public health resources.

We introduce a model with an underlying mathematical structure, incorporating a cross interaction parameter, and conduct numerical simulations to assess the extent of the spread under different scenarios of integration or segregation. Additionally, we examine the impact of implementing non-medicinal interventions aimed at reducing the infection’s extent, comparing the outcomes with those of the original case without interventions.

The manuscript is organized as follows: After the introduction in section (1), we present a novel two-population, two-reservoir model in section (2), with different levels of segregation and integration. We derive the basic reproductive number and introduce the effects of non-medicinal interventions. Our results and numerical simualtions are discussed in section (3) before concluding in section (4).

## 2 Theoretical Framework

### 2.1 A two-population, two-reservoir model

We introduce a novel compartmental model with two interacting populations, to simulate Cholera spreads in societies with different degrees of integration or segregation. This is an SIBR model, where each population is divided into four compartments, *S* (susceptible), *I* (infected), *B* (concentration of the bacteria in infected water reservoirs), and *R* (removed). We neglect the transmission from person to person and focus solely on transmission through exposure to infected water reservoirs.

We present our model in system (1). The susceptible compartments are populated through natural births and depopulated through natural deaths and cholera infections. We denote by *N*_*i*_ the total number of individuals in population *i*, by *µ*_*i*_ the natural death rate or birth rate of population i, which assume to be equal, without loss of generality. The rate of infection depends on the susceptible population *S*_*i*_, the rate of exposure to infected water resources *β*_*i*_, the human infectious dose *κ* and the reservoir bacteria concentration *B*. The human infectious dose is defined such that when *κ* = *B*, the probability of infection upon exposure to the reservoir is 50%.

The infectious compartment is augmented by infections from susceptibles defined above and it is reduced by the natural death rate and by the removal rate *γ* (due to recovery or disease induced death). We ignore infections caused by direct human to human interaction, as this would be a marginal factor in comparison with the main driving force of infection: bacterial contamination of vital environmental sources like water.

In the bacteria reservoirs *B*_*i*_, the deposition of bacteria from the infected class of a population to its corresponding reservoir happens at a rate of *χ*_*i*_ while the deposition rate into the other reservoir occurs at *τ χ*_*i*_. The parameter *τ* is a suppression parameter that represents the cross interaction between between a population with the other natural reservoir, and could be used to represent the degree of segregation or integration between the populations, with numerical values 0 ≤ *τ* ≤ 1. Finally, the rate of death of the Vibrio cholerae bacteria in the environment is given *δ*.

The recovery compartments *R*_*i*_ occur due the removal rate *γ*_*i*_ (by recovery or disease related death) from the infected population. It is diminished by natural deaths among the surviving recovered population only, since a fraction *l* of those removed are already dead due to disease.

The flow diagram in Fig. (1) summarizes the dynamics of the model among its various compartments, interactions and their rates. The red lines in this figure represent the dynamics of infection of susceptible individuals upon contact with the corresponding contaminated reservoir, the black lines connecting *I*_*i*_ and *R*_*i*_ summarize the rate of recovery of infectious individuals and the dashed blue lines represent the deposition of the bacteria by infectious individuals into their own as well as the other reservoirs. The other one-sided black lines show the natural birth and death rates.

**Figure 1:**
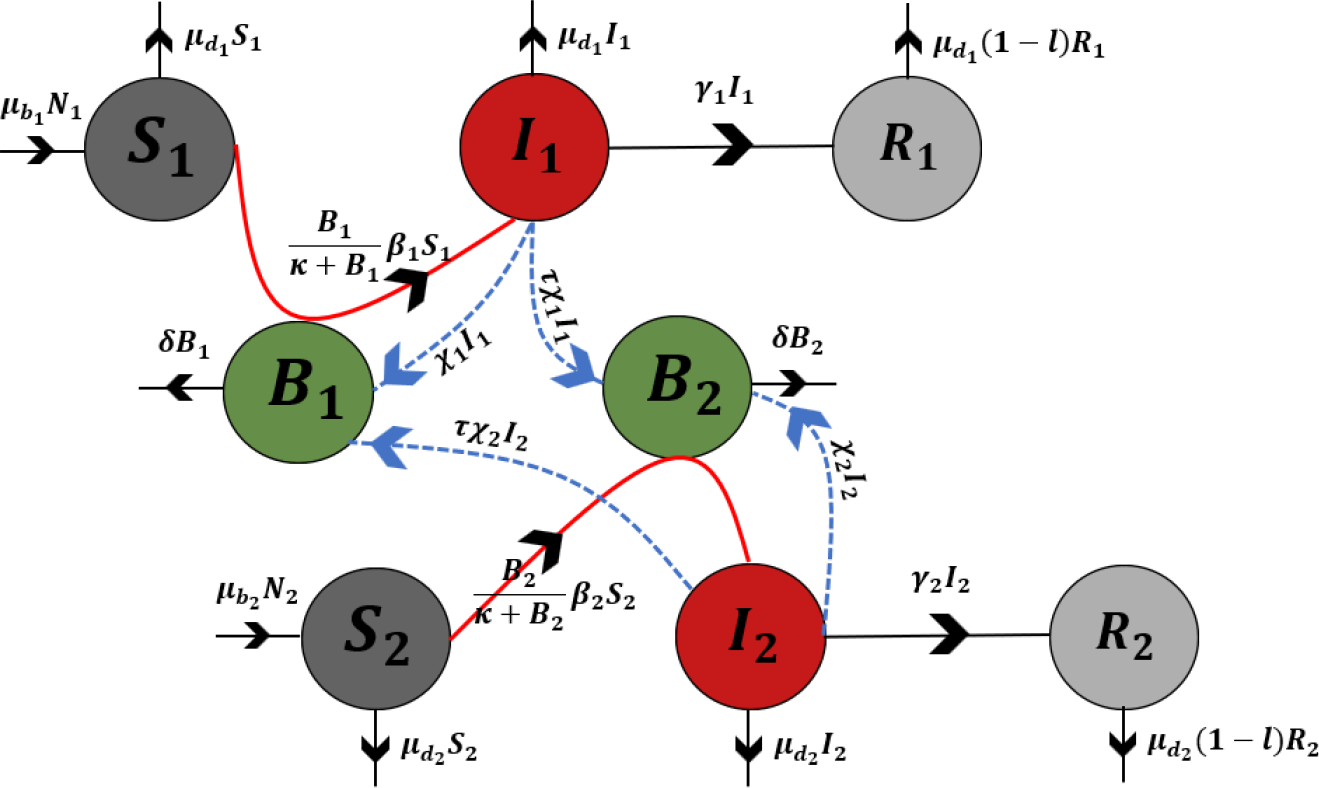
A flow diagram corresponding to the two-population two-reservoir model presented in system (1). The flow lines represent dynamics between compartments and the dashed lines correspond to deposition into the reservoirs.

**Table 1:** A summary of the model parameters and their definitions.

| Parameter | Description |
| --- | --- |
| $\mu_{b_i}$ | Rates of natural birth |
| $\mu_{d_i}$ | Rates of natural death |
| $\beta_i$ | Rates of contact/expousre to infected resources |
| $\kappa$ | Human infectious dose - 50% infection rate upon exposure |
| $\gamma_i$ | Rates of removal (recovery or death) from Cholera infection |
| $\chi_i$ | Rate of bacterial deposition from infected people to their reservoir |
| $\tau$ | Supression factor representing levels of segregation or integration |
| $l$ | Rate of death among infectious individuals |
| $\delta$ | Rate of death of the Vibrio Cholerae bacteria in the reservoir |

Mathematically, our model is best represented by the following system of coupled ordinary differential equations:

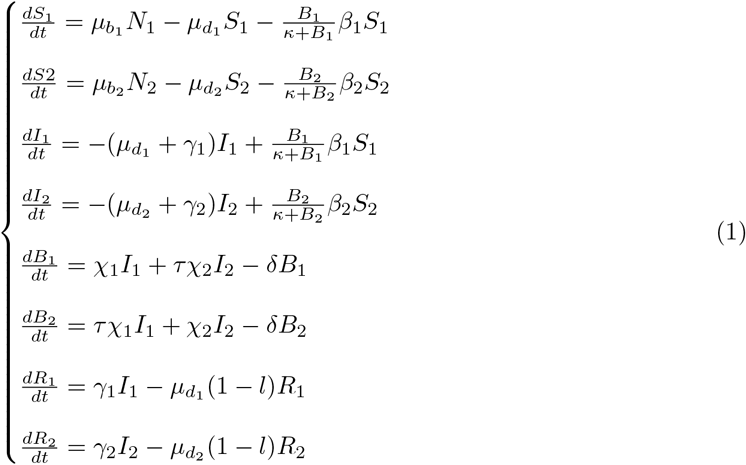

where *N*_*i*_ = *S*_*i*_ + *I*_*i*_ + *R*_*i*_. An important quantity in epidemic model is the force of infection which is essential rate for determining infections. For Cholera spreads, the force of infection is mainly caused by the interaction between the susceptibles and the bacteria in the reservoir. In this model, it is given by:

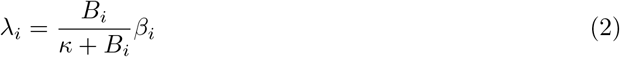

#### Lemma 1

*λ* is an increasing function with respect to *β* and B and decreasing with respect to *κ*.

**Proof:** We can verify that: 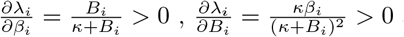and 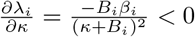.

Higher concentrations of bacteria in the reservoir and higher contact rates result in a stronger rate of infection, while a higher human infectious dose means less rate of infections.

Without loss of generality, we assume here that the rates of natural deaths and natural births are equal, hence: 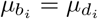. The net change in population would consequently be:

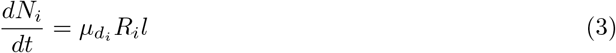

which represents the net decrease in the population due to deaths caused by cholera infections. Note that in this setting, the total populations *N*_*i*_ decrease due to disease induced deaths and equal natural birth and death rates.

#### Theorem 1: Fully segregated populations are independent

For *τ* = 0 the two populations are independent.

**Proof:** For *τ* = 0, the equations for *B*_*i*_ in system (1) decouple and the interaction between two populations is fully suppressed. Consequently the whole system decouples into two independent systems given by:

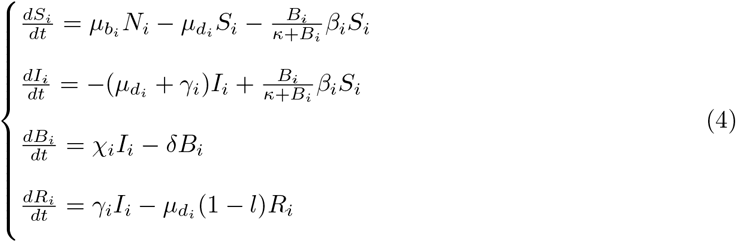

for *i* = 1, 2.

#### Theorem 2 Fully integrated populations form a single population

For *τ* = 1, two fully integrated populations form a single population which SIR compartments are the sums of the individual compartments.

**Proof:** Two fully integrated populations would have access to same reservoirs, environmental and social factors and contact rates, hence we assume that each doublet of parameters 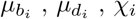, *χ*_*i*_ and *B*_*i*_ to be identical. In addition, with equal access to both reservoirs, we have that *τ* = 1.

Under these conditions, the model can be reduced to describe the spread of cholera in one population.

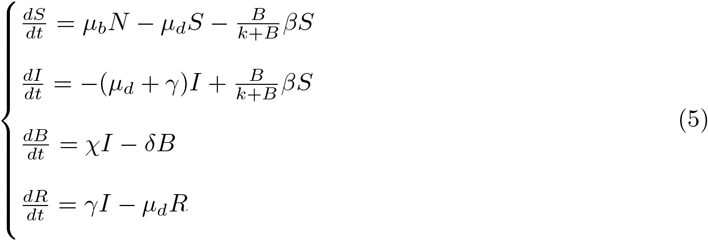

where *S* = *S*_1_ + *S*_2_, *I* = *I*_1_ + *I*_2_, *R* = *R*_1_ + *R*_2_ and *N* = *N*_1_ + *N*_2_

In this paper, we consider a set of realistic scenarios of two interacting populations under various levels of segregation or integration, modelled by 0 ≤ *τ* ≤ 1.

### 2.2 Basic reproductive number

The model presented above admits a disease-free equilibrium given by

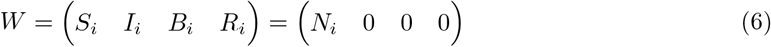

This equilibrium is asymptotically stable if the basic reproductive number *R*_0_ is less than one. To determine the basic reproductive number we use the next generation matrix approach. First, let us define 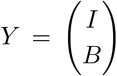 where *I* = *I*_1_ + *I*_2_ and *B* = *B*_1_ + *B*_2_ Let us now consider the infected classes of the first population 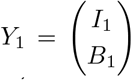 and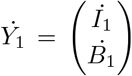. We then divide 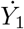 into *f − v* such that 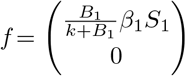 and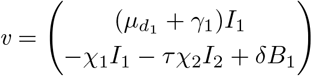. The Jacobian of f and *v* evaluated at the disease-free equilibrium are respectively F and V.

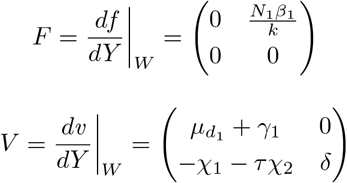

The next generation matrix is given by

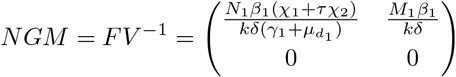

The basic reproductive number is given by the dominant eigenvalue of the next-generation matrix.

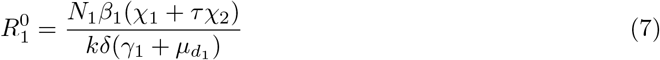

Similarly, we can show that

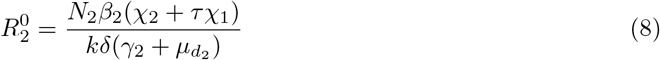

#### Lemma 2

The basic reproductive number of the two populations are related through the cross interaction parameter *τ*

**Proof:** Dividing both sides of the expressions in equations (7) and (8), we get that:

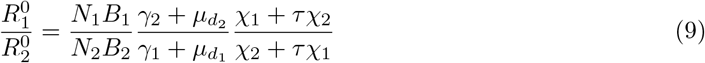

From eq. (9), setting the constant parameter as 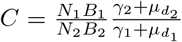 and the *τ* dependent part as 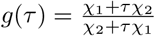, we could express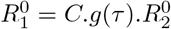.

### 2.3 Non-medicinal interventions

Various non-medicinal strategies can be employed to prohibit the transmission of cholera, such as ensuring access to clean water, promoting sanitation, and emphasizing personal hygiene. Implementing hygiene practices such as the promotion of handwashing, proper sanitation infrastructure, and access to clean water significantly contributes to breaking the cycle of cholera transmission. Educational campaigns on safe water storage and food handling further enhance community resilience. Additionally, community engagement and the establishment of effective waste disposal systems are essential in preventing the contamination of water sources. Non-medicinal interventions not only address the immediate challenges posed by cholera but also contribute to long-term public health improvements, creating a foundation for sustainable disease prevention and control.

The impact of these interventions is reflected in the dynamic model’s mathematical parameters [39], with no alterations to the model’s structure itself. Sanitation measures, for instance, help prevent water contamination from human feces by separating it from the drinking water supply, thereby reducing contamination rates represented by *χ*_*i*_. Chlorinating water reduces bacterial content and increases the removal rate *δ*, while purifying drinking water through boiling or filtering lowers bacterial concentrations denoted as *B*_*i*_. Additionally, introducing alternative sources of clean drinking water diminishes the interaction between susceptible individuals and contaminated water, thereby lowering the contact rate *β*_*i*_ [42, 43]. It is assumed that a combination of these measures will be implemented, varying in scale within different populations, and the model’s outcomes are re-evaluated by considering the effects of these changes on model parameters.

We presume in this paper that non-medicinal interventions due to public health campaigns and local initiatives would affect both populations leading to a realistic decrease by 20% and 50% in the contamination rates and contact rates respectively among both populations as well as an increase of 50% in the bacterial removal rates. We implement the adjusted parameters due to these interventions in our two-reservoir two-population model to forecast the expected subsequent reduction in the number of infections.

## 3 Results, numerical simulations and discussions

Fig. (2) presents 3 three-dimensional plots of *λ* in Eq. (2) in terms of *B* and *β, κ* and *B*, and *β* and *κ* respectively (using *Wolfram MATHEMATICA 13*.*3*). Our numerical plots are shown for *κ* = 10^5^ cells/mL in (a), *β* = 0.07 /day in (b) and for *B* = 10^6^ cells/mL in (c). The 3D plots confirm the outcomes of Lemma 1. We can clearly see that *λ* increases with respect to *B* and *β* in (a), increases with *B* and decreases with *κ* in (b), and finally increases with *β* and decreases with *κ* in (c).

**Figure 2:**
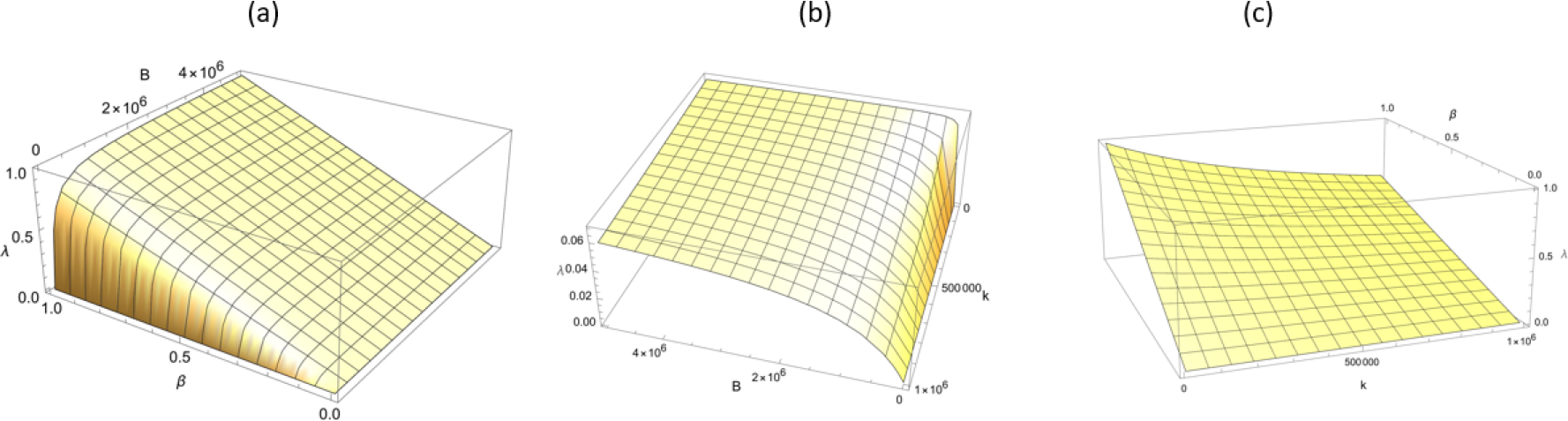
Three dimensional plots of the force of infection *λ* in terms of: (a) *B* and *β* on the left, (b) *κ* and *B* in the middle and (c) *β* and *κ* on the right.

The parameters used in our numerical analysis are based on values listed and used in the available literature [36, 39]. Before non-medicinal interventions, medications and vaccines, we assume that the contact rate between susceptibles and their reservoirs are *β*_1_ = 0.015 /day and *β*_2_ = 0.04 /day.

The recovery rates among the infectious are 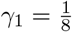 / day and 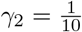 / day. The contributions of the infectious to the reservoir vibrio concentration are *χ*_1_ = 2 cells/mL/day and *χ*_2_ = 10 cells/mL/day while the half saturation rate (the concentration of bacteria that leads to a 50% infection rate) is *κ* = 10^5^ cells/mL. The natural birth/death rates are given by 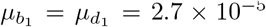 / day and 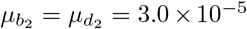 / day while the percentage of deaths caused by Cholera among the infectious individuals is *l* = 4%. The population sizes are given by a sample of *N*_1_ = *N*_2_ = 10^5^ individuals. The suppression parameter of cross interaction *τ* which accounts for various levels of segregation or integration between the two populations is varied between 0 *≤ τ ≤* 1. All the subsequent simulations corresponding to Figs. (3-7) were performed in *MATLAB R2023b*.

**Figure 3:**
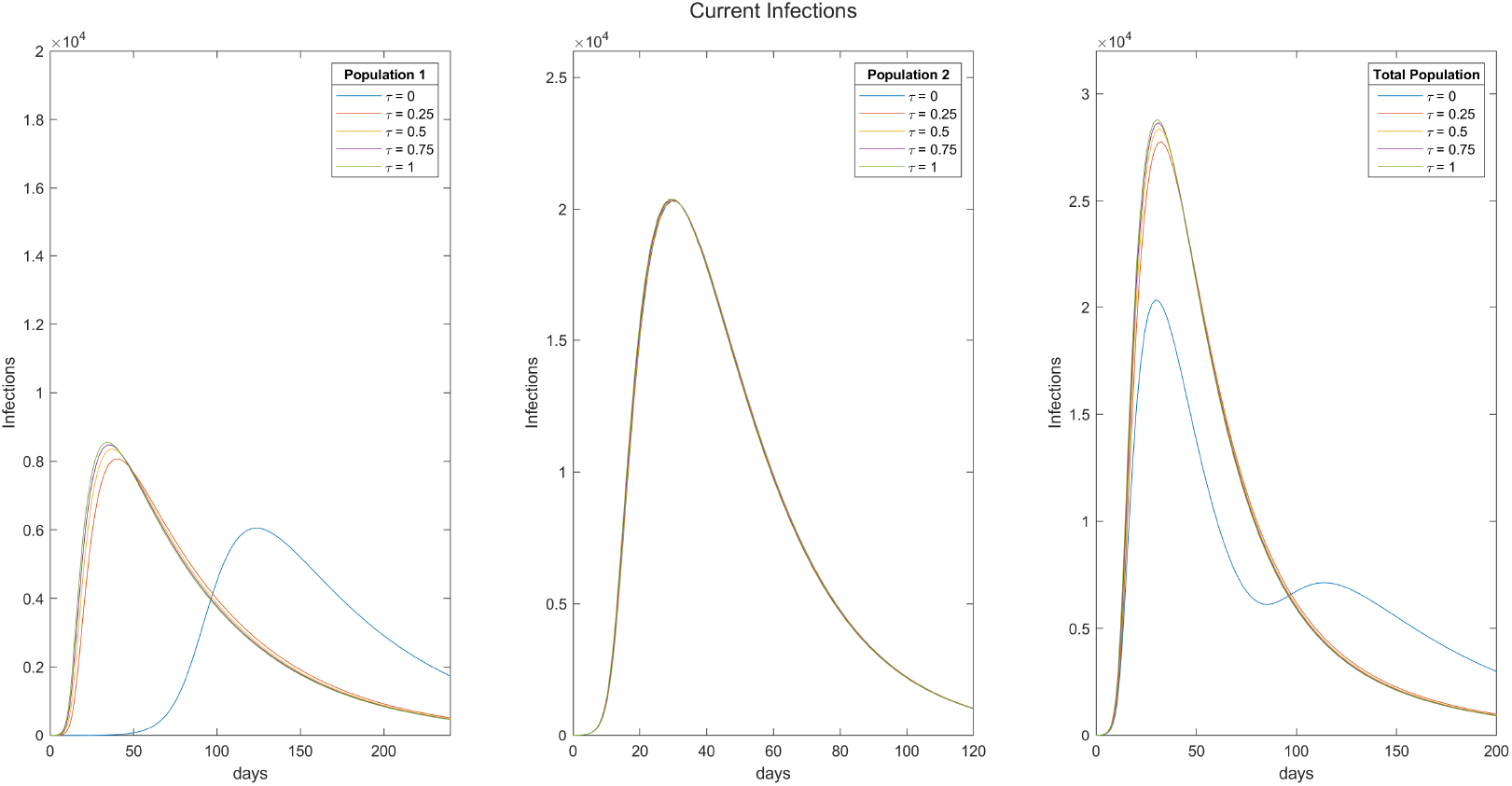
The current number of infections in population 1 on the left, population 2 in the middle and the total population on the right for various values of *τ* between 0 and 1.

**Figure 4:**
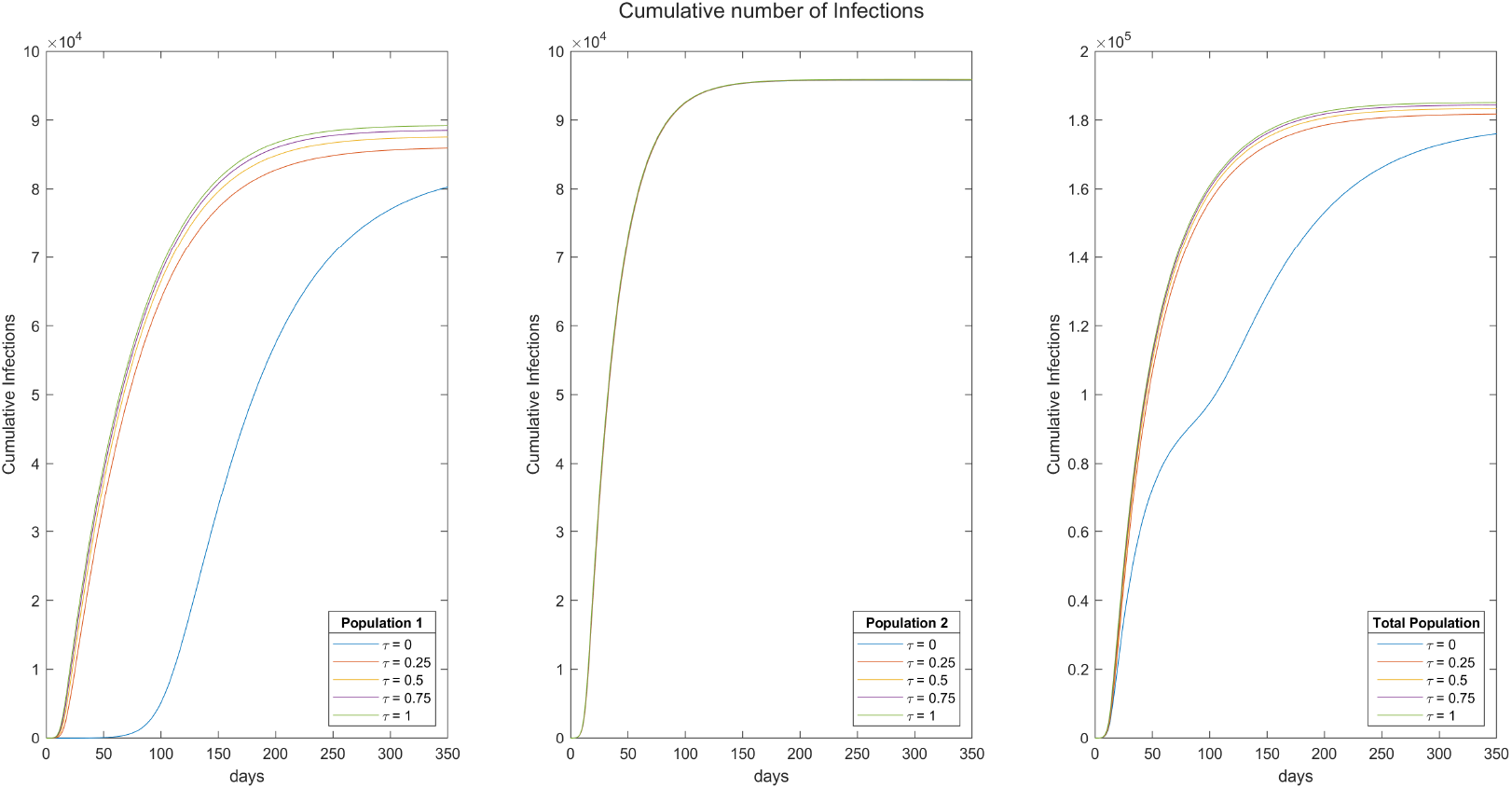
The cumulative number of infections in population 1 on the left, population 2 in the middle and the total population on the right for various values of *τ* between 0 and 1.

**Figure 5:**
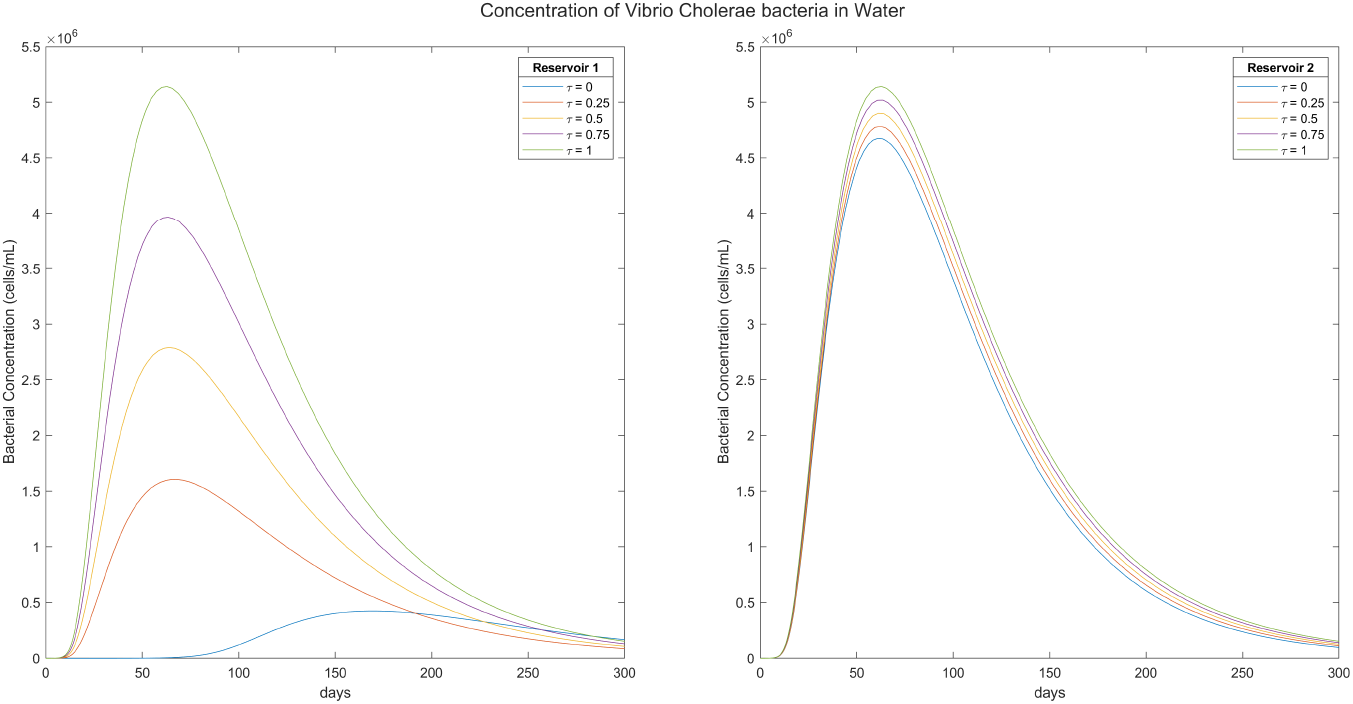
The concentration of the Vibrio Cholerae bacteria in water in the environmental reservoirs 1 and 2 belonging to populations 1 and 2 for various levels of the cross interaction parameter *τ* with values between 0 and 1.

**Figure 6:**
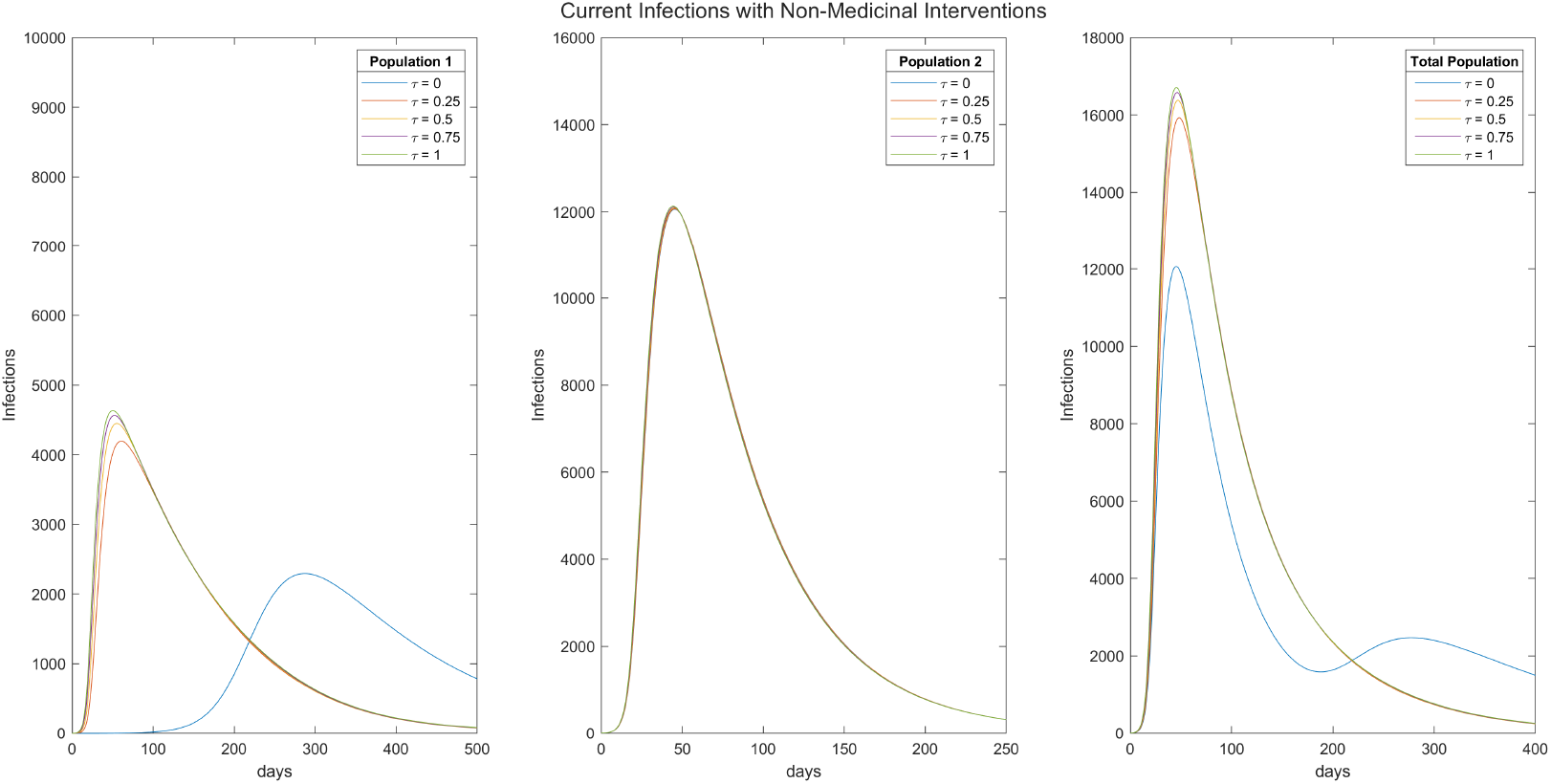
The current number of infections in population 1 on the left, population 2 in the middle and the total population on the right for various values of *τ* upon the implementation of non-medicinal interventions.

**Figure 7:**
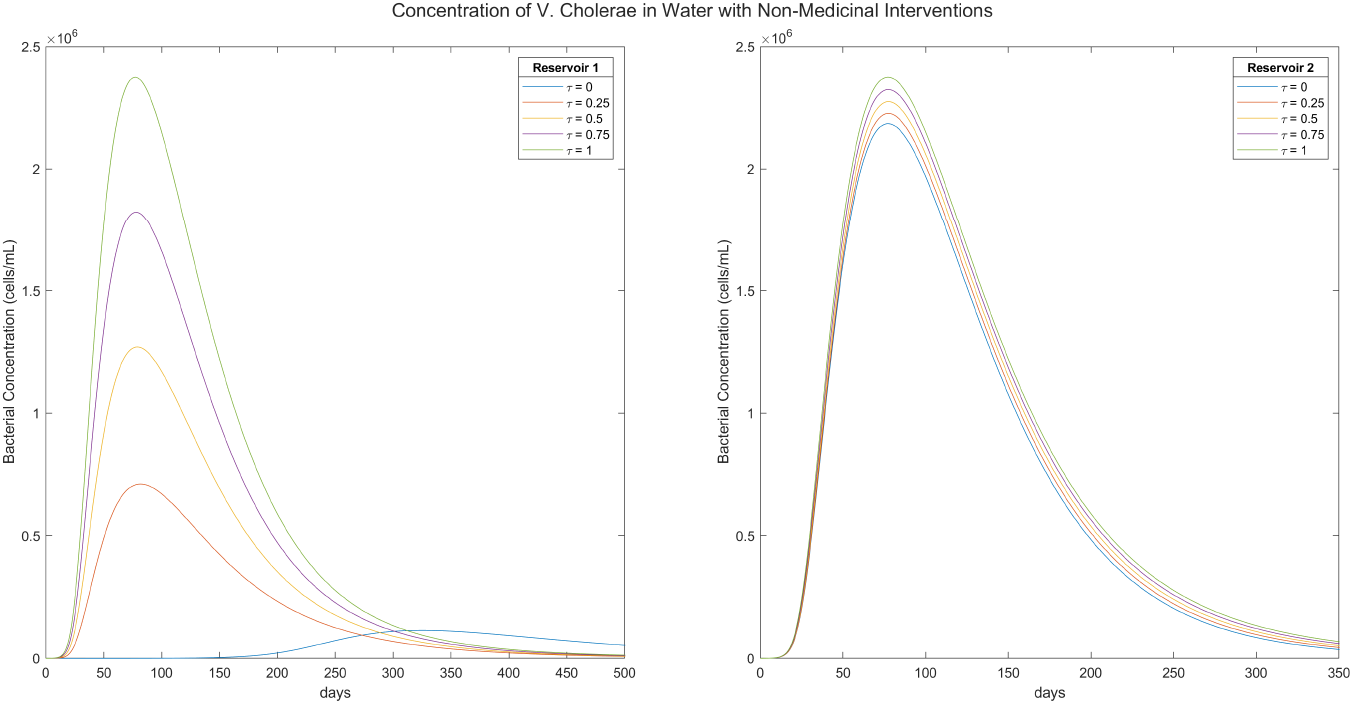
The concentration of the Vibrio Cholerae bacteria in water in the environmental reservoirs 1 and 2 belonging to populations 1 and 2 after the implementation of non-medicinal interventions for various levels of the cross interaction parameter *τ*.

In Fig. 3, we forecast the anticipated number of infections in the absence of interventions across various levels of segregation or integration, represented by the suppression factor *τ* = 0, 0.25, 0.5, 0.75 and 1, This gradient reflects different degrees of population interaction, ranging from complete segregation to full integration. Our assumption is that the conditions for infection growth are more favorable in population 2. Our findings indicate that the daily number of infections in population 2 (middle plot) is minimally impacted by its interaction with reservoir 1. Conversely, interaction with the other reservoir significantly amplifies infections in population 1. The plot on the left, displaying the blue curve, illustrates that the lowest endemic occurs when population 1 is entirely isolated from population 2. In addition to reducing infections (and subsequently hospitalizations and deaths), this scenario also results in a delay of approximately 100 days in the peak of the spread. This delay provides authorities with more time to implement public health and preventive measures. The right plot illustrates the total daily infections stemming from both populations. Isolating the two populations from each other’s reservoirs (*τ* = 0) clearly decreases the total number of infections, alleviating about a third of infections and their associated hospitalizations and deaths. While most of the reduction in infections is observed in population 1, a more controlled total spread ensures additional resources are available for the sick in population 2 benefiting both communities mutually. Notably, a secondary peak in total infections corresponds to a late rise in infections in population 1. However, this peak occurs at a significantly delayed time, with a weaker amplitude, and can be mitigated through vaccination and medicinal intervention, given the available time before its emergence.

The cumulative number of anticipated infections is depicted in Fig. 4 for population 1 (left), population 2 (middle), and the total population (right) across values of *τ* ranging from 0 to 1. Consistently, we observe a reduction in the cumulative number of infections in population 1 (left) with decreasing interaction with the other population. The sharp increase occurs at a significantly delayed time when *τ* = 0, indicating complete isolation from the other community. Similarly, the cumulative number of infections in population 2, as shown in the middle plot, is not notably affected by the level of interaction, given its role as the host of stronger endemic conditions. Consequently, isolating individuals from the corresponding population’s environmental reservoir results in the lowest total number of infections in the overall population, as depicted in the right plot. There is a considerable time delay before reaching the plateau corresponding to the peak of cumulative infections. When considered alongside the findings from Fig. 3, our results affirm the significance of complete isolation of individuals from the environmental resources of others to effectively contain the spread and mitigate the disease.

The concentration of the Vibrio Cholerae bacteria in water in the environmental reservoirs 1 and 2 belonging to populations 1 and 2 respectively are displayed in Fig. 5 for different levels of cross interaction represented by 0 ≤ *τ* ≤ 1. Our simulation shows that the bacteria concentration is highly impacted by cross interaction. For population 1, suppressing the cross interaction completely can reduce the magnitude of peak concentration by more than ten folds. In addition, the peak concentration could be delayed up to 15 weeks for maximum segregation of the two populations, giving public health authorities ample time to intervene and mitigate the related disease. Even for intermediate levels of suppression of *τ*, the peak concentration could be reduced by several orders of magnitude in the reservoir of population 1. These important gains are mutual for both communities, as the peak concentration of bacteria would also be reduced in reservoir 2, but in a less significant fashion. Our forecasts show that various levels of suppression of cross interaction would decrease the peak of Vibrio Cholerae presence in the second reservoir by up to 15% in the best case scenario of maximal separation of the two communities. Intermediate levels of cross interaction would lead to less decreases in population 2. This result shows that curbing interactions across communities and their corresponding reservoirs during Cholerae disease spreads would significantly decrease peak bacteria levels in their reservoirs. In line to our analytical result in Lemma 1 and our numerical result in Fig. 2, a decrease in bacteria concentration *B* ultimately leads to a decrease in the force of infection *λ*, hence to a a diminished number infections *I*.

We expect that upon the spread of Cholera, related public health authorities would intervene and start by taking preliminary measures that aim to mitigate the disease through non-medicinal interventions before other medicinal and vaccination measures could be taken. We inspect the anticipated changes that would be achieved through a reduction of 20% in the contamination rates *χ*_*i*_ and of 50% in contact rates *β*_*i*_ respectively among both populations as well as an increase of 50% in the bacterial removal rate *δ*.

We present that outcomes of our simulation of these reductions on the number of current infections in Fig. 6, corresponding to the same population modelled in Fig. 3. We find that the number of infections is highly reduced in both population and for all levels of cross community interactions, and consequently in the total number of infections. We can see that the peak of infections fall about 40% in each population for every value of *τ*. Moreover, for population 1 (left), the peak for scenarios involving some degree of cross immunity interactions is delayed by about 6 weeks, while that of total suppression of interactions (*τ* = 0) is delayed by more than 20 weeks, practically allowing the public health administration enough time to implement all additional medicinal and vaccination measures needed. In population 2 (middle), we can also observe an average of a 6 week delay in maximal number of infections, with a similar observation regarding extents of reduction and peak delays among the total population (right). In this best case scenario of full separation between the two communities and each other’s reservoirs, accompanied by a timely application of non-medicinal interventions, the Cholera endemic could be largely controlled and mitigated, reducing the threat that it would constitute to pubic health.

We finally forecast the effect of implementation of non-medicinal interventions with their previously anticipated effects on deposition rates, contact rates and bacterial removal rate numerically. We find that bacterial concentrations would be more than halved due to these measures in both environmental reservoirs for all levels of interactions between the communities. In addition, peak concentrations would be delayed by about 5 *−* 6 weeks in most scenarios, as we can see by comparing the outcomes plotted in Fig. 7 to those in Fig. 5 corresponding to no interventions. In particular, for a full segregation scenario, reservoir 1 bacterial concentrations would be slashed by more than four times and their peak concentrations would occur by more than 25 weeks later (left). The reservoir of population 2 undergoes similar reductions in the magnitude of peak contamination and delays in its occurrence except for the *τ* = 0 case, which doesn’t cause a severe reduction as in the case of reservoir 1 (right). Nevertheless, it still sees a reduction of more than 50% of its value without these non-medicinal interventions.

## 4 Conclusions

In this paper, we introduced a dynamic compartmental model tailored to portray Cholera outbreaks within a dual-population framework, incorporating two environmental reservoirs. The model aimed to simulate the influence of segregation or integration between two populations on disease transmission, considering both non-medicinal interventions and scenarios without such measures.

Analytically, we demonstrated the direct dependence of the force of infection of each population on the contact rates and contamination rates, and its inverse dependence on the human infectious dose. We also showed that fully segregated populations are independent while fully integrated populations could be treated as a single population, and that the reproductive numbers of the two populations are related through a proportionality function of the suppression parameter.

By permitting infectious individuals to engage with the reservoir of the alternate population at varying suppression levels alongside their regular interaction with their own reservoir, our simulations revealed that heightened suppression of cross-community interaction effectively reduced overall infection rates and those in the less contaminated and contacted population. Furthermore, we observed substantial delays in the onset of peak infections, granting crucial time for public health interventions and that decreased cross-immunity interactions were also associated with a decline in bacterial concentrations in environmental reservoirs.

Ultimately, our numerical analysis demonstrated that non-medicinal interventions, encompassing sanitation and water purification, played a pivotal role in significantly reducing and delaying infections. This affords concerned parties a valuable timeframe for the implementation of additional medicinal measures, underscoring the potential impact of integrated public health strategies in mitigating Cholera outbreaks.

## Data Availability

No data has been used in this study

